# Hepatitis B and D: a forecast on actions needed to reduce incidence and achieve elimination

**DOI:** 10.1101/2021.08.11.21261940

**Authors:** Scott Greenhalgh, Andrew Klug

## Abstract

**Background:** Viral hepatitis negatively affects the health of millions, with the worst health outcomes associated with the hepatitis D virus (HDV). Fortunately, HDV is rare and requires prior infection with the hepatitis B virus (HBV) before it can establish infection and transmit. As such, public health officials have opted to indirectly control HDV by reducing HBV incidence, primarily through hepatitis B vaccination, which has dramatically reduced HBV incidence since its rollout in the 1980s. However, investigations into the consequences of hepatitis B vaccination on both the control and evolution of HDV remain limited.

**Methods:** We developed a mathematical model of HBV and HDV transmission to investigate the effects of hepatitis B vaccination on both HBV and HDV. We calibrated our model to the HBV and HDV transmission scenarios occurring in Sub-Saharan Africa, and estimate HBV vaccination thresholds that cause HBV and HDV elimination, inhibit the spread of virulent HDV strains, and achieve the targets set by public health authorities for reducing all viral hepatitis incidence by 90%.

**Results:** Our findings illustrate hepatitis B vaccination rates above 0.0096 year^−1^ and 0.018 year^−1^ will likely achieve HDV and HBV elimination, respectively. Furthermore, in the majority of transmission settings, scaling-up vaccination rates to at least 0.009 year^−1^ or 0.02136 year^−1^ will achieve the 90% reduction in hepatitis D and B, respectively, called for by health authorities.

**Conclusion:** Our results suggest a scale-up of hepatitis B vaccination is required to achieve the targets set by public health officials for reducing all viral hepatitis by 90%. Furthermore, the scale-up required to achieve such targets would bring HBV and HDV to the brink of the vaccination threshold required for their elimination. Thus, with sufficient investments to scale up global hepatitis vaccination, prevention, testing, and treatment services, the eradication of both HBV and HDV are likely feasible endeavors, especially once the 90% reduction goal is reached.

## 1. Introduction

Hepatitis viruses are estimated to infect 2.3 billion individuals worldwide [1]. Of these individuals, the 12.4 million [2,3] who are chronically co-infected with the hepatitis B (HBV) and hepatitis D (HDV) viruses likely face the worst prognosis, as the odds of liver cancer are more likely, and the progression towards liver-related death occurs much more rapidly [3]. Fortunately, two key facts offer hope against these negative outcomes. First, HDV infection is uncommon, as it requires prior infection with HBV to infect and transmit [3]. Second, the annual incidence of HBV continues to plummet in many regions due to the successful rollout of highly efficacious HBV vaccines. In fact, the HBV vaccine has been so successful that it has been adopted into 184 government immunization programs [2], which have been a driving force in its continued ability to reduce the global prevalence of HBV.

While the effect of the HBV vaccine on HBV incidence and prevalence are well-studied [4–6], far less is known about its impact on the transmission of HDV. In part, this lack of information is because some do not consider HDV to be a relevant medical problem [7]. It is this stance that once led to a decline in screening for HDV infection [8,9], and its subsequent rebound in parts of Europe [7]. Even with the rebound of HDV, some public health officials continue to only control HDV indirectly through sustained efforts to reduce HBV, such as HBV vaccination, the use of harm reduction services, and education on both blood and injection safety [3], instead of investing time and resources into developing HDV specific health interventions. Regardless of whether specific health interventions should be developed for HDV, there are still important questions that remain unanswered concerning HDV’s status in the world. For instance, it is unknown what level of HBV vaccination is required so the decline in current HDV incidence reaches the reductions called for by the Global Health Sector Strategy on Viral Hepatitis 2016-2021 [10]. Similarly, it is also unknown whether the current HBV vaccination levels are sufficiently high to cause the elimination of HDV, as ongoing surveillance illustrates a marked decline of HDV in the Western world, among other regions [7].

To inform on the potential for HBV vaccination to aid in meeting the targets of the Global Health Sector Strategy on Viral Hepatitis 2016-2021 [10], its potential to cause HDV elimination, and its ability to inhibit the transmission of virulent HDV strains, we developed a mathematical model of HBV and HDV transmission. Our mathematical model considers the most common transmission routes for both HBV and HDV infection, namely person-to-person transmission, in addition to acute and chronic stages of infection. To calibrate our mathematical model, we used freely available data on HBV and HDV [7,11–13], as well as demographic data from regions that feature low, intermediate, and high HDV prevalence, along with vaccination coverages of 13.4%, 24.7%, and 62.1%, respectively [14]. We also estimated the vaccination levels required to reduce HBV and HDV incidence by 90% (one of the primary objectives of the Global Health Sector Strategy on Viral Hepatitis 2016-2021), the levels required to cause both HBV and HDV elimination, and the levels required to inhibit the spread of virulent HDV strains.

## 2. Materials and Methods

To evaluate the effect of the HBV vaccination on inhibiting the spread and virulence evolution of HDV, we created a mathematical model of HBV and HDV transmission (Webappendix). Through the mathematical model, we estimate the effect that HBV vaccination has on HDV transmission by reducing the number of individuals who are susceptible to HDV infection. We consider scenarios where HBV vaccination achieves 13.4%, 24.7%, and 62.1% coverage [14], with vaccine efficacies of 80%, 90%, or 100% [15] to measure the associated decline in HDV incidence, identify the vaccination required to reduce hepatitis incidence by 90%, and determine the thresholds that cause hepatitis B and D elimination. In addition, to provide further credibility on the merit of HBV vaccination for controlling the spread and evolution of HDV, we also evaluate the potential spread of mutant strains of HDV, and the impact that this might have on the control and elimination of both HBV and HDV.

### 2.1 Mathematical model

For our mathematical model, we consider a population divided into classes based on disease status. For HBV, this amounts to individuals susceptible to infection (*S*), acute HBV infected individuals (*A*_*B*_), chronic HBV infected individuals (*C*_*B*_), individuals recovered from HBV infection (*R*_*B*_), and HBV vaccinated individuals (*V*). For HDV, due to the requirement of HBV infection, we have acute HBV and HDV infected individuals ([*A*_*B*_*A*_*D*_]), chronic HBV and acute HDV infected individuals ([*C*_*B*_*A*_*D*_]), chronic HBV and HDV infected individuals ([*C*_*B*_*C*_*D*_]) and individuals with chronic HBV infection that have recovered from HDV ([*C*_*B*_*R*_*D*_]). Our model considers the rate that susceptible individuals become infected with HBV to be

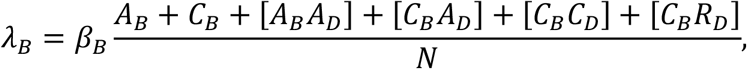

where *β*_*B*_ is the transmission rate of HBV and *N* is the total population. In addition, the rate that HBV infected individuals become co-infected with HDV is

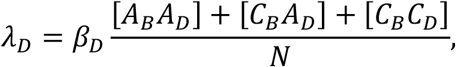

where *β*_*D*_ is the transmission rate of HDV.

Also considered in our model, is the birth rate (*b*), the proportion of newborns with hepatitis (*σ*), the rate latent hepatitis infection clears or becomes active (*γ*), the proportion of latent hepatitis infections that lead to active disease (*ρ*), the recovery rate from hepatitis infection (*δ*), the hepatitis associated death rate (*μ*), and the vaccination rate for HBV (*ν*). Note, parameter subscripts of *B* and *D* correspond to the parameters associated with HBV infection and HDV infection, respectively. Further details of parameters, including values and sources, are available in Table 1.

**Table 1.** Parameter values and sources.

| Parameter | Variable | Value | Distribution | Source |
| --- | --- | --- | --- | --- |
| Population size (billions) | $N$ | 1.08 | | [21] |
| Birth rate | $b$ | $0.024 \text{ year}^{-1}$ | | [26] |
| Death rates |  |  |  |  |
| Natural | $\mu$ | $1/78.8 \text{ year}^{-1}$ | $\text{Exp}(1/78.8)$ | [27] |
| HBV | $\mu_B$ | $0.00086 \text{ year}^{-1}$ | $\text{Exp}(0.00086)$ | Webappendix [28] |
| HDV | $\mu_D$ | $0.0030 \text{ year}^{-1}$ | $\text{Exp}(0.0030)$ | Webappendix [29] |
| Proportion of newborns (from infected parents) with |  |  |  |  |
| chronic HBV | $\sigma_B$ | 0.2 | $U[0.0,0.4]$ | [30] |
| chronic HDV | $\sigma_D$ | $0.05 \cdot \sigma_B$ | | [31] |
| Transmission rate |  |  |  |  |
| HBV | $\beta_B$ | $0.391 \text{ year}^{-1}$ | | Webappendix |
| HDV | $\beta_D$ | $16.45 \text{ year}^{-1}$ | | Webappendix |
| Incubation period |  |  |  |  |
| HBV | $1/\gamma_B$ | 91.25 days | $\text{Tri}[45,68.75,160]$ | [32] |
| HDV | $1/\gamma_D$ | 91.25 days | $\text{Tri}[45,68.75,160]$ | [32] |
| Proportion that develop active disease |  |  |  |  |
| HBV | $\rho_B$ | 0.1 | | Webappendix [28] |
| HDV | $\rho_D$ | 0.1 | | Webappendix [28] |
| Average duration of chronic infection with HBV | $1/\delta_B$ | 50 years | $\delta_B \sim \text{Exp}(1 / 50)$ | [20] |
| Average duration of chronic infection with HDV | $1/\delta_D$ | 50 years | $\delta_D \sim \text{Exp}(1 / 50)$ | [20] |
| Vaccination rate for HBV | $\nu$ | | $U[0,0.026]$ | Webappendix |

### 2.2 The reproductive numbers of HBV and HDV

To provide insight on the potential for the HBV vaccine to eradicate both HBV and HDV we estimated the basic [16,17], control [18], and effective [18] reproductive numbers. The reproductive numbers of HBV are estimated using the next-generation method (Webappendix). Using this approach, we have that the effective reproductive number is calculated as

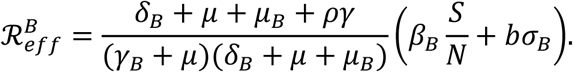

Evaluating the effective reproductive number at the disease-free equilibrium, we obtain the control reproductive number for HBV:

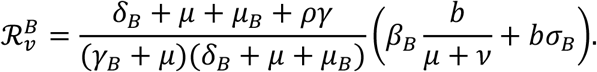

Further imposing that *ν* = 0, we obtain the basic reproductive number for HBV:

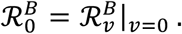

From the control reproductive number, we determine the critical HBV vaccination level to cause elimination by solving 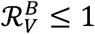 for *ν*.

With regards to HDV, we also use the next-generation method to estimate the reproductive numbers (Webappendix). Using this approach, we have that the control reproductive number for HDV is

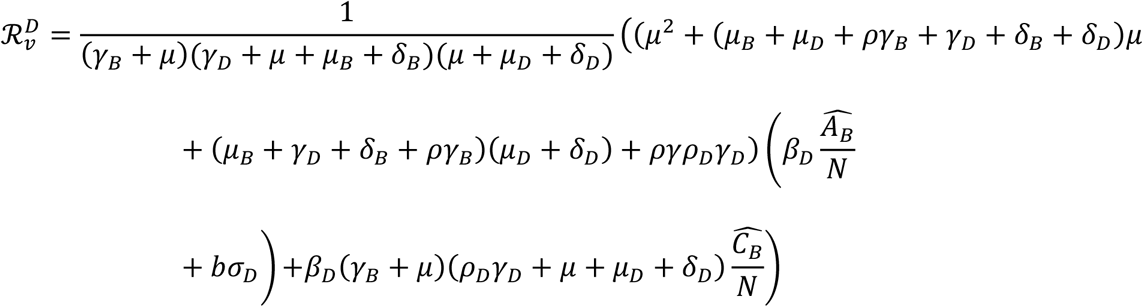

where 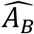 and 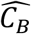 correspond to the HBV endemic and HDV free equilibrium values (Webappendix). It naturally follows that the basic reproductive number is

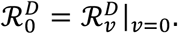

From 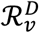, the critical HBV vaccination level to cause the elimination of HDV is determined by solving 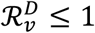 for *ν*.

### 2.3 Invasion analysis

To determine whether mutant HDV strains can usurp the currently circulating HDV strain, we consider an extension of our HBV and HDV transmission model to include an additional mutant HDV strain (Webappendix). Using this extended system, we examine the potential for the mutant HDV strain to invade by the use of the mutant HDV strain’s basic reproductive number:

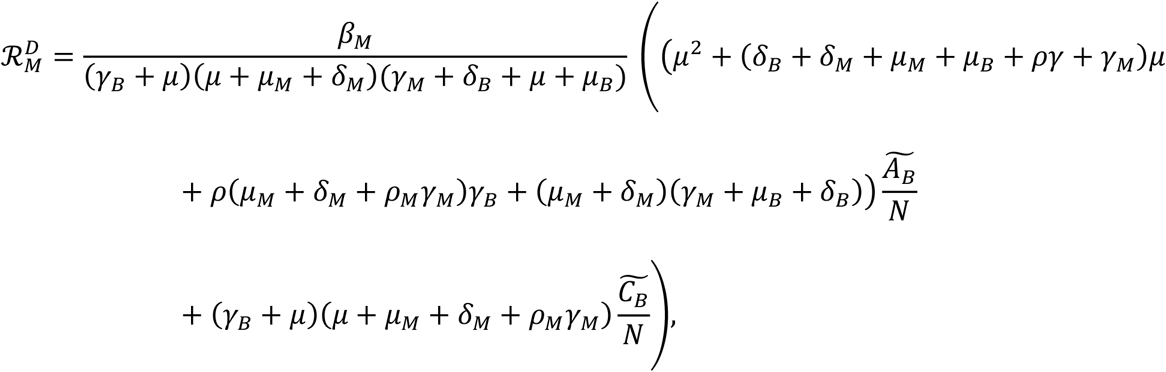

where 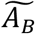 and 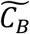 are the values of the acute and chronically infected HBV individuals at the HBV and HDV co-endemic equilibrium (Webappendix).

By the competitive exclusion principle [19] a mutant disease strain is out-competed by a resident strain that features a higher basic reproductive number. So, we determine the HBV vaccination level so that 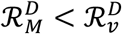 for mutant HDV strains that feature enhanced transmission rates *β*_*M*_ that are 0.75, 1.0, 1.25, 1.50, or 1.75-fold that of *β*_*D*_. We also evaluate the effect of enhanced incubation periods and average durations of chronic infection on the overall ability for mutant HDV strains to invade.

### 2.4 Transmission scenarios

To capture the effect of HBV vaccination on HBV and HDV incidence we consider scenarios of high, intermediate, and low HBV transmission intensity, and base HBV and HDV transmission intensity on estimates of their basic reproductive numbers [11,20]. In addition, we base model demographics on the population of Sub-Saharan Africa [21] due to the variety of HBV and HDV transmission settings that occur in the region.

#### HBV transmission and vaccination

We consider baseline scenarios to be when HBV vaccination rates achieve 13.4%, 24.7%, and 62.1% coverage [14] with vaccine efficacies of 80%, 90%, and 100% [15]. We classify these scenarios as high, intermediate, and low endemic HBV regions, respectively, and obtain two estimates of the required increase in HBV vaccination rates needed to achieve a 90% reduction in HBV incidence. The first estimate using the HBV endemic and HDV free equilibrium to determine the required increase in HBV vaccination rate, while the second uses the HBV and HDV co-endemic equilibrium. In addition, to calibrate HBV transmission rates, we use estimates of the basic reproductive number for HBV [11], and estimates of HBV recovery and mortality rates (Table 1, Webappendix).

#### HDV transmission

To determine the transmission intensity of HDV, we use estimates of its reproductive number from the literature [20]. Furthermore, we estimate the required increase in HBV vaccination rates to achieve a 90% reduction in HDV incidence under high, intermediate, and low HBV transmission intensities, using the HBV and HDV co-endemic equilibrium.

### 2.5 Sensitivity analysis

To quantify how each parameters’ uncertainty contributes to the variability in predictions of control reproductive numbers, we calculated variance-based first-order sensitivity indices [22]. Details of the probability distributions used in the calculation of indices are available in Table 1.

## 3. Results

To determine the effect of HBV vaccination on the control, elimination, and virulence evolution of HDV we simulated the transmission of HBV and HDV while calibrated to HBV vaccination rates that achieve 13.4%, 24.7%, and 62.1%vaccination coverage, respectively [14]. Using these baseline values, we estimated the required increase in vaccination to reduce HBV and HDV incidence by 90%, the vaccination thresholds that eliminate HDV alone, in addition to both HBV and HDV, along with the vaccination levels required to inhibit the invasion of more virulent HDV strains under high, intermediate, and low endemic HBV settings. We gauged the risk for HDV evolution by considering mutant strains with transmission rates of 75%, 100%, 125%, 150%, and 175% the resident HDV strain transmission rate.

In the absence of HDV, we found that the vaccination rates required to achieve the vaccination coverages of 13.4%, 24.7%, and 62.1% are 0.0033 year^−1^, 0.0068 year^−1^, and 0.019 year^−1^ for high, intermediate, and low HBV transmission intensities, respectively, at the HBV endemic and HDV free equilibrium. Using these rates as baseline values and the HBV endemic and HDV free equilibrium, a 90% reduction in HBV incidence requires increasing the vaccination rates to 0.0166 year^−1^, and 0.0170 year^−1^ for the high and intermediate transmission intensities, respectively (Figure 1a). For the low transmission intensity, the vaccination rate of 0.019 year^−1^ causes 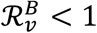 (Figure 2), thereby rendering the endemic equilibrium infeasible. When considering both HBV and HDV transmission at the co-endemic equilibrium, we found that the vaccination rates required to achieve HBV vaccination coverages were similar to those of the HBV endemic and HDV free equilibrium, namely 0.0033 year^−1^, 0.0068 year^−1^, and 0.023 year^−1^, respectively. However, achieving a 90% reduction in HBV incidence required further increasing the HBV vaccination rates to 0.021 year^−1^, and 0.0237 year^−1^ (Figure 1b), for the high and intermediate transmission intensities, respectively. Our results also show to achieve a 90% reduction in HDV incidence in regions with high or intermediate levels of HBV transmission intensity requires a lower increase in the HBV vaccination rate, namely HBV vaccination rates of 0.0089 year^−1^, and 0.009 year^−1^.

**Figure 1.**
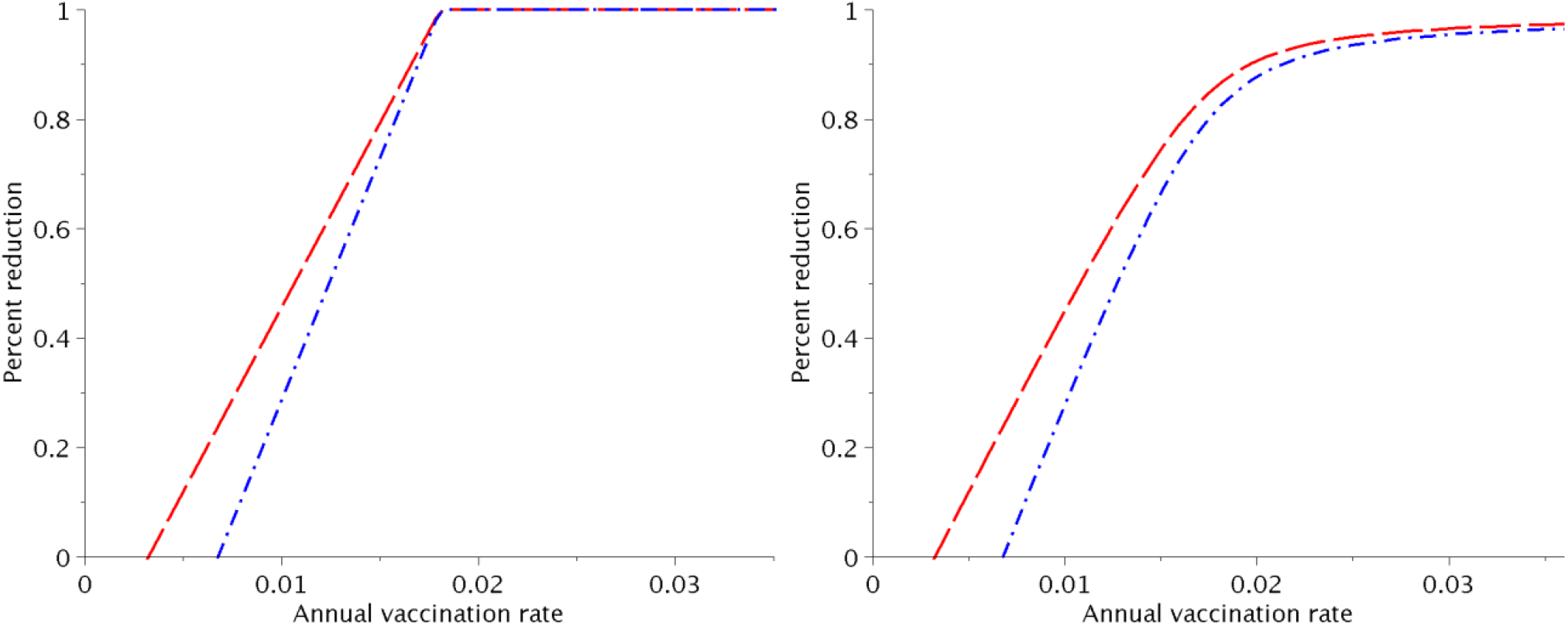
Percent reduction in incidence for given vaccination rate. a) Reduction at HBV endemic and HDV free equilibrium for high (red dashed line) and intermediate (blue dash-dot line) transmission, and b) Reduction at HBV and HDV co-endemic equilibrium for high (red dashed line), and intermediate (blue dash-dot line) transmission.

**Figure 2.**
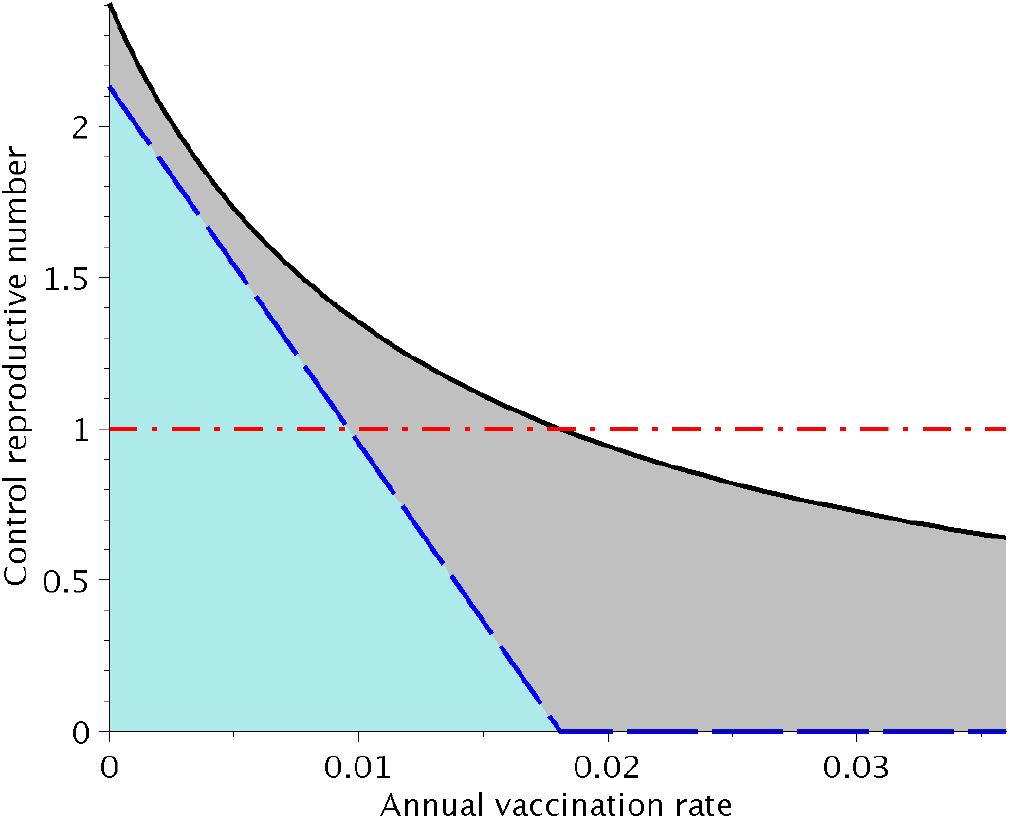
Control reproductive number for HBV and HDV versus the annual vaccination rate. The critical value for disease elimination (red dash-dot line), 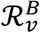 (black solid line) and 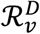 (blue dashed line).

Our model predicts increasing HBV vaccination rates to levels that cause a 90% reduction in HBV and HDV incidence would avert 2.64 (95% CI 2.54, 2.73), and 2.04 (95% CI 1.96, 2.12), and HBV incidence per 1000 people over 5 years, under high and intermediate HBV transmission intensity (Figure 3). These HBV vaccination rates would also prevent 0.151 (95% CI 0.138, 0.163), and 0.113 (95% CI 0.103, 0.123) HDV incidence per 1000 people over 5 years, under the same HBV vaccination scenarios. The HBV incidence averted increases to 9.66 (95% CI 9.31, 10.00), and 7.44 (95% CI 7.15, 7.74), and 0.679 (95% CI 0.629, 0.729) per 1000 people, and 0.510 (95% CI 0.469, 0.550) per 1000 people for HDV incidence if a 10-year period is considered (Figure 3).

**Figure 3.**
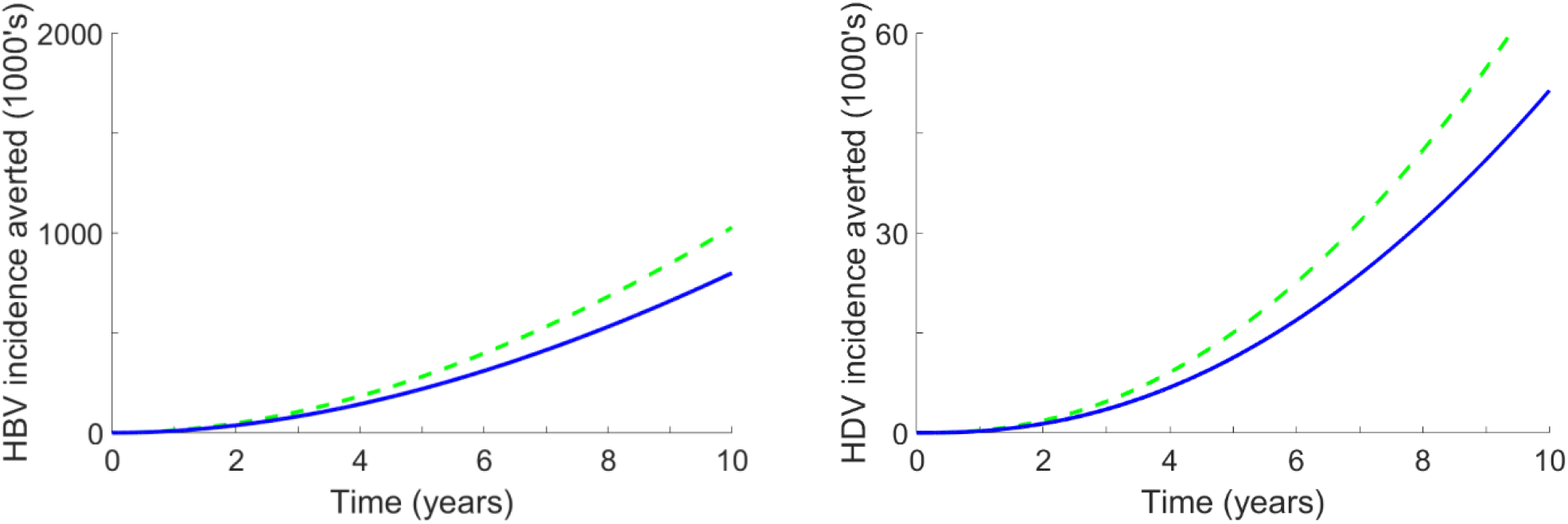
HBV and HDV incidence averted. Incidence averted was calculated by subtracting mean HBV and mean HDV trajectories for high (green dashed line), and intermediate (blue solid line) settings under HBV vaccination rates that cause a 90% reduction in HBV incidence from incidence predicted with baseline HBV vaccination rates. Mean trajectories are calculated from model predictions using 1000 random parameter samples.

Our results indicate mutant strains of HDV are unlikely to usurp currently circulating HDV strains. Specifically, for an HBV vaccination rate of at least 0.0033 year^−1^, our results show that a mutant strain of HDV would need to feature a transmission rate over 125% that of the currently circulating HDV strain and at least a 1.8-fold longer incubation period, or at least a 1.2-fold longer average duration of chronic infection (Figure 4, Figure 5).

**Figure 4.**
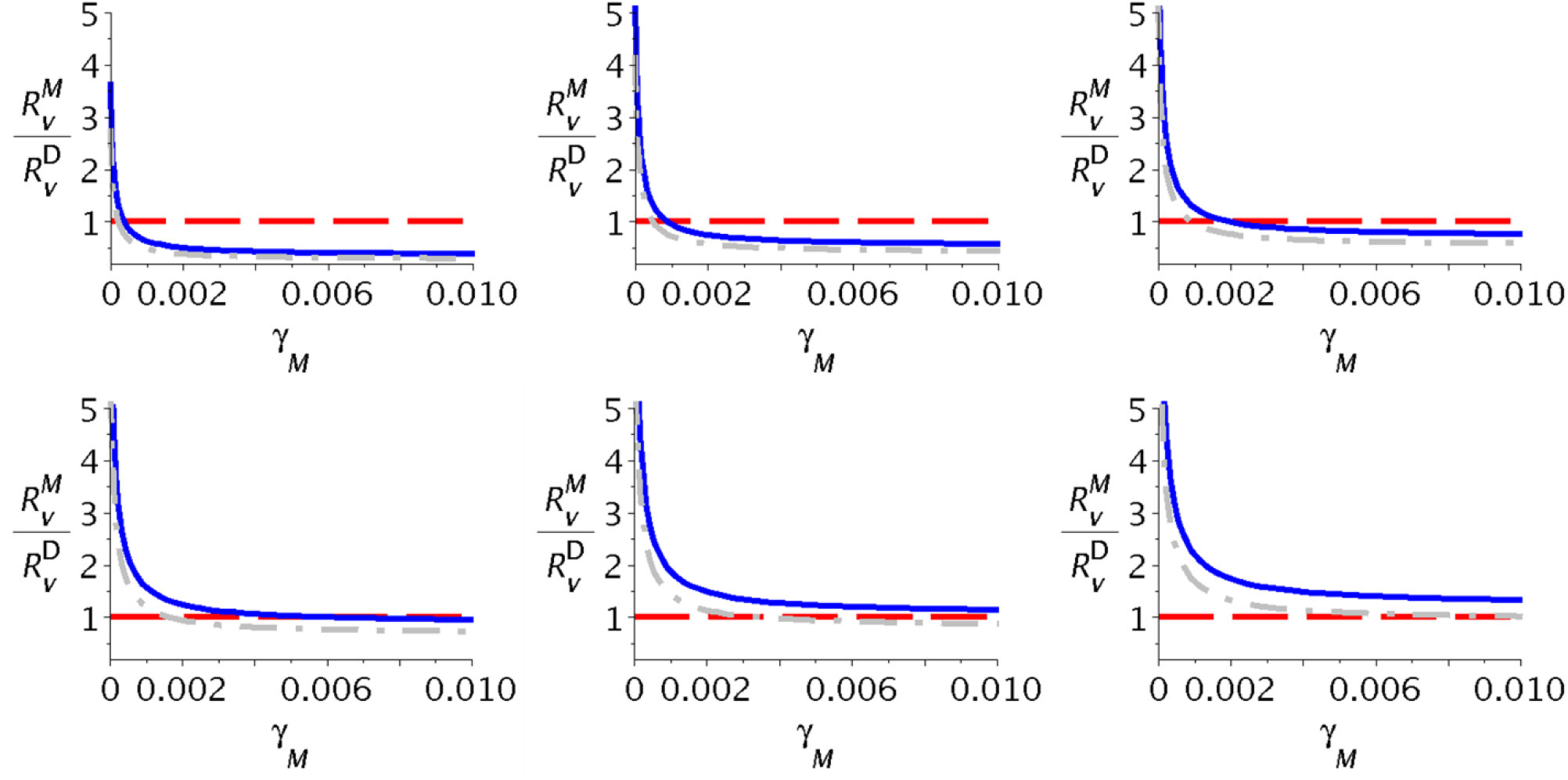
The ratio of control reproductive numbers versus incubation rate. The ratio features a mutant HDV strain with a) *β*_*M*_ = 0.5*β*_*D*_, b) *β*_*M*_ = 0.75*β*_*D*_, c) *β*_*M*_ = 1.0*β*_*D*_, d) *β*_*M*_ = 1.25*β*_*D*_, e) *β*_*M*_ = 1.5*β*_*D*_, and f) *β*_*M*_ = 1.75*β*_*D*_ along with vaccination rates *ν* of 0.0068 (grey dash-dot line), and 0.0033 (blue solid line) per year, respectively, with the critical value for invasion of 1 (red dashed line).

**Figure 5.**
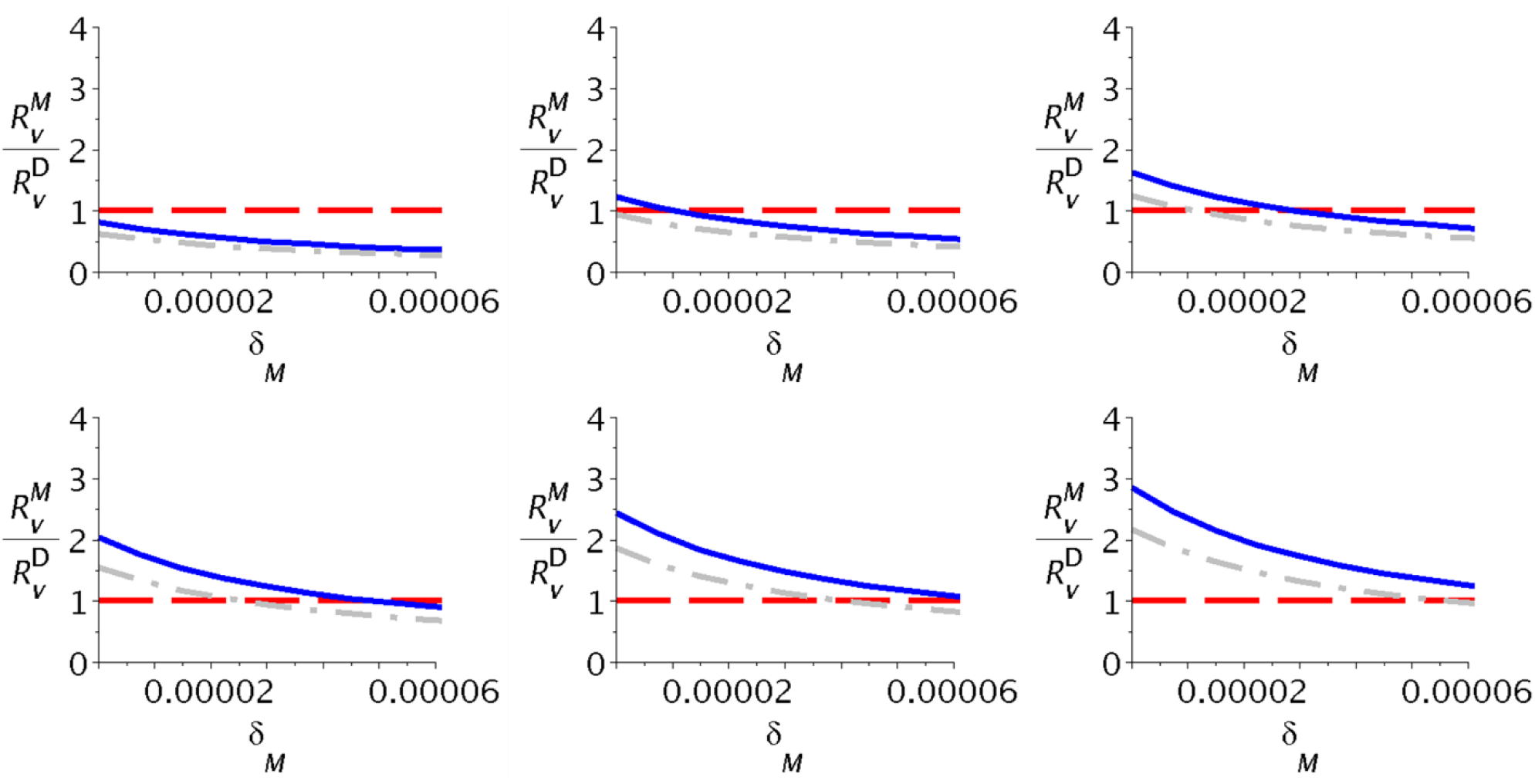
The ratio of control reproductive numbers versus the average duration of chronic infection for the mutant HDV strain. The ratio features a mutant HDV strain with a) *β*_*M*_ = 0.5*β*_*D*_, b) *β*_*M*_ = 0.75*β*_*D*_, c) *β*_*M*_ = 1.0*β*_*D*_, d) *β*_*M*_ = 1.25*β*_*D*_, e) *β*_*M*_ = 1.5*β*_*D*_, and f) *β*_*M*_ = 1.75*β*_*D*_ along with vaccination rates *ν* of 0.0068 (grey dash-dot line), and 0.0033 (blue solid line) per year, respectively, with the critical value for invasion of 1 (red dashed line).

The sensitivity analysis of model parameters shows that 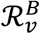 and 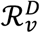 were most sensitive to the natural death rate and the duration of chronic infection with HBV (Figure 5). Beyond these parameters, 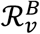 was also sensitive to the HBV vaccination rate, although this had little effect 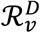(Figure 5). In a similar fashion, 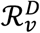 was sensitive to the duration of chronic infection with HDV, but this parameter had a negligible impact on 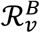 (Figure 6). Other model parameters had a minimal impact on any variability of both 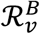 and 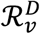 (Figure 6).

**Figure 6.**
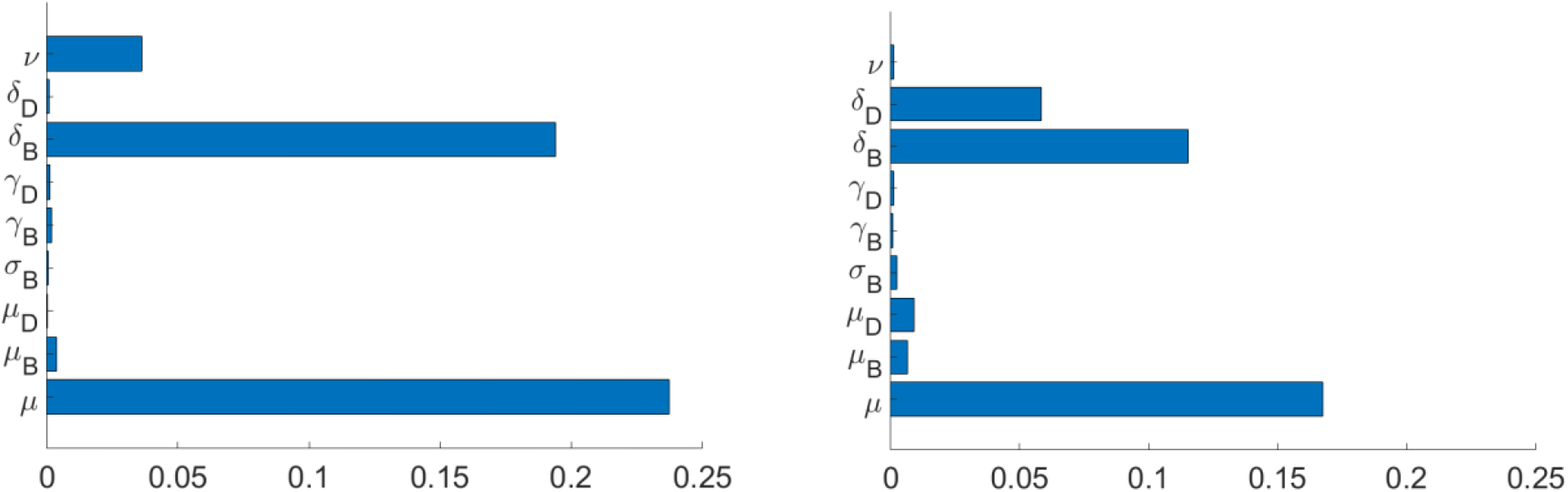
Variance-based sensitivity indices. a) control reproductive number for HBV and b) control reproductive number for HDV. Index calculation is based on 10,000 random parameter samples.

## 4. Discussion

The analysis of our mathematical model of HBV and HDV coinfection predicts that HBV vaccination levels beyond 0.018 year^−1^ will eliminate both HBV and HDV. For moderate HBV vaccination levels, our model shows that while there are risks associated with mutant HDV strains, a dramatic increase in disease transmissibility, incubation period, or duration of chronic infection would need to occur to offset the benefit of vaccination.

The findings of our work reaffirm theoretical studies that show HDV has an impact on the spread and control of HBV [20]. Specifically, our results indicate that the presence of HDV requires a higher HBV vaccination rate to cause the elimination of both diseases. A potential workaround of this issue is to first focus on HDV elimination, given the relatively low HBV vaccination rate required to accomplish this feat, and then focus on HBV elimination. While such an endeavor would likely take a longer period to complete, it would entail less upscaling of HBV vaccination and therefore may be more feasible in regions where health resources are scarce.

As our work considers HBV and HDV transmission among a general population, an important direction for future work is to include demographic structure to reflect the role that at-risk communities, such as drug injectors and sex workers, play in enabling persistence of both HBV and HDV. Similarly, modifications to our model to reflect individuals’ knowledge of HBV and HDV infection status and stratification by age would also likely prove to be fruitful avenues for further investigation.

To obtain our results, we made several simplifying assumptions. To begin, our model only accounts for stages of acute and chronic infection for both HBV and HDV. Therefore, our model does not reflect other disease stages, such as superinfection, latent infection, or the difference between chronically infected individuals that receive or do not receive drug treatment, such as entecavir for HBV [23], or in-development drug treatments for HDV [24]. We also did not account for the impact of coinfection with other types of viral hepatitis that commonly occur, such as hepatitis C [25]. In addition, our model is calibrated to describe HBV and HDV transmission in Sub-Saharan Africa, although with modest modifications it could be used to evaluate the HBV and HDV interventions under different demographic settings.

## Conclusion

In summary, our results indicate that HDV elimination is likely underway in the vast majority of low HBV transmission settings. Our results also suggest that modest increases in HBV vaccination rates in moderate and high HBV transmission settings would lead to the elimination of HBV and HDV. As such, our results reinforce the idea that the eradication of HBV and HDV are feasible endeavors. Thus, with sufficient investments to scale up global hepatitis prevention, testing, and treatment services, both HBV and HDV could be eradication in time for the WHO’s 2030 target date.

## Data Availability

All data generated or analyzed during this study are included in this published article and its supplementary information files.

## List of abbreviations

HBV: Hepatitis B virus
HDV: Hepatitis D virus
WHO: World Health Organization

## Declarations

### Ethics approval and consent to participate

Not applicable.

### Consent for publication

Not applicable.

### Competing interests

The authors declare that they have no competing interests.

### Funding

Funding support for AK was provided by Siena College’s Center for Undergraduate Research and Creative Activity.

### Author’s Contributions

SG came up with the study design, AK and SG contributed to model development and analysis. All authors participated in the drafting and editing of the manuscript.

## Notes

### Competing Interest Statement

The authors have declared no competing interest.

### Funding Statement

AK was supported by Siena College's Center for Undergraduate Research and Creative Activity

## References

1. Jefferies M, Rauff B, Rashid H, Lam T, Rafiq S. Update on global epidemiology of viral hepatitis and preventive strategies. World J Clin cases. 2018;6: 589–599. doi:10.12998/wjcc.v6.i13.589

2. Nelson NP, Easterbrook PJ, McMahon BJ. Epidemiology of Hepatitis B Virus Infection and Impact of Vaccination on Disease. Clin Liver Dis. 2016;20: 607–628. doi:10.1016/j.cld.2016.06.006

3. WHO. Hepatitis D Factsheet [Internet]. Geneva; 2019. Available: https://www.who.int/news-room/fact-sheets/detail/hepatitis-d

4. Bruxvoort K, Slezak J, Huang R, Sy LS, Towner W, Ackerson B, et al. 286. Hepatitis B Vaccine Compliance: Comparing 2-Dose and 3-Dose Vaccines. Open Forum Infect Dis. 2019;6: S156--S156. doi:10.1093/ofid/ofz360.361

5. Nayagam S, Thursz M, Sicuri E, Conteh L, Wiktor S, Low-Beer D, et al. Requirements for global elimination of hepatitis B: a modelling study. Lancet Infect Dis. 2016;16: 1399–1408. doi:10.1016/S1473-3099(16)30204-3

6. Razavi-Shearer D, Gamkrelidze I, Nguyen MH, Chen D-S, Van Damme P, Abbas Z, et al. Global prevalence, treatment, and prevention of hepatitis B virus infection in 2016: a modelling study. Lancet Gastroenterol Hepatol. 2018;3: 383–403. doi:10.1016/S2468-1253(18)30056-6

7. Rizzetto M. Hepatitis D Virus: Introduction and Epidemiology. Cold Spring Harb Perspect Med. 2015;5: a021576. doi:10.1101/cshperspect.a021576

8. Cross TJS, Rizzi P, Horner M, Jolly A, Hussain MJ, Smith HM, et al. The increasing prevalence of hepatitis delta virus (HDV) infection in South London. J Med Virol. 2008;80: 277–282. doi:10.1002/jmv.21078

9. Holmberg SD, Ward JW. Hepatitis Delta: Seek and Ye Shall Find. J Infect Dis. 2010;202: 822–824. doi:10.1086/655809

10. WHO. Global Health Sector Strategy on Viral Hepatitis 2016-2021 [Internet]. Geneva; 2016. Available: https://apps.who.int/iris/bitstream/handle/10665/246177/WHO-HIV-2016.06-eng.pdf

11. Zou L, Zhang W, Ruan S. Modeling the transmission dynamics and control of hepatitis B virus in China. J Theor Biol. 2010;262: 330–338. doi:10.1016/j.jtbi.2009.09.035

12. Kim WR. Epidemiology of hepatitis B in the United States. Hepatology. 2009;49: S28--34. doi:10.1002/hep.22975

13. Chen H-Y, Shen D-T, Ji D-Z, Han P-C, Zhang W-M, Ma J-F, et al. Prevalence and burden of hepatitis D virus infection in the global population: a systematic review and meta-analysis. Gut. 2018; doi:10.1136/gutjnl-2018-316601

14. Mei-Chuan Hung, Walter W. Williams, Peng-Jun Lu, David K. Kim, Lisa A. Grohskopf, Tamara Pilishvili, Tami H. Skoff, Noele P. Nelson, Rafael Harpaz, Lauri E. Markowitz, Alfonso Rodriguez-Lainz APF. Vaccination Coverage Among Adults in the United States, National Health Interview Survey, 2016. 2018.

15. Hamborsky J, Kroger A, Wolfe S. Epidemiology and prevention of vaccine-preventable diseases [Internet]. 3th ed. Washington, D.C.: Center for Disease Control and Prevention; 2015. Available: https://www.cdc.gov/vaccines/pubs/pinkbook/downloads/table-of-contents.pdf

16. Heffernan JM, Smith RJ, Wahl LM. Perspectives on the basic reproductive ratio. J R Soc Interface. 2005;2: 281–293. doi:10.1098/rsif.2005.0042

17. van den Driessche P, Watmough J. Reproduction numbers and sub-threshold endemic equilibria for compartmental models of disease transmission. Mathematical Biosciences. 2002. pp. 29–48. doi:10.1016/S0025-5564(02)00108-6

18. van den Driessche P. Reproduction numbers of infectious disease models. Infect Dis Model. 2017;2: 288–303. doi:10.1016/j.idm.2017.06.002

19. Martcheva M. An Introduction to Mathematical Epidemiology [Internet]. Boston, MA: Springer US; 2015. doi:10.1007/978-1-4899-7612-3

20. Xiridou M, Borkent-Raven B, Hulshof J, Wallinga J. How hepatitis D virus can hinder the control of hepatitis B virus. PLoS One. 2009;4: e5247. doi:10.1371/journal.pone.0005247

21. World Bank. Population, total. 2018.

22. Sobol IM. Global sensitivity indices for nonlinear mathematical models and their Monte Carlo estimates. Math Comput Simul. 2001;55: 271–280. doi:10.1016/S0378-4754(00)00270-6

23. WHO. Hepatitis B Factsheet [Internet]. Geneva; 2018. Available: https://www.who.int/news-room/fact-sheets/detail/hepatitis-b

24. Mentha N, Clément S, Negro F, Alfaiate D. A review on hepatitis D: From virology to new therapies. J Adv Res. 2019;17: 3–15. doi:10.1016/j.jare.2019.03.009

25. Caccamo G, Saffioti F, Raimondo G. Hepatitis B virus and hepatitis C virus dual infection. World J Gastroenterol. 2014;20: 14559–14567. doi:10.3748/wjg.v20.i40.14559

26. US Department of Commerce. 2007-2011 American Community Survey 5-Year Estimates [Internet]. US Census Bureau; 2016. Available: https://www.census.gov/programs-surveys/acs/technical-documentation/table-and-geography-changes/2011/5-year.html

27. National Center for Health Statistics. Health, United States, 2015: With Special Feature on Racial and Ethnic Health Disparities. [Internet]. Hyattsville, Md; 2016. Available: https://www.cdc.gov/nchs/data/hus/hus15.pdf

28. Wang T. Model of life expectancy of chronic hepatitis B carriers in an endemic region. J Epidemiol. 2009;19: 311–318. doi:10.2188/jea.je20090039

29. Negro F. Hepatitis D Virus Coinfection and Superinfection. Cold Spring Harb Perspect Med. 2014;4: a021550.–a021550. doi:10.1101/cshperspect.a021550

30. Yelemkoure ET, Yonli AT, Montesano C, Ouattara AK, Diarra B, Zohoncon TM, et al. Prevention of mother-to-child transmission of hepatitis B virus in Burkina Faso: Screening, vaccination and evaluation of post-vaccination antibodies against hepatitis B surface antigen in newborns. J Public Health Africa. 2018;9: 816. doi:10.4081/jphia.2018.816

31. WHO. Global hepatitis report [Internet]. 2017. Available: https://www.who.int/hepatitis/publications/global-hepatitis-report2017/en/

32. Goyal A, Murray JM. Recognizing the impact of endemic hepatitis D virus on hepatitis B virus eradication. Theor Popul Biol. 2016;112: 60–69. doi:10.1016/j.tpb.2016.08.004

33. Otto SP, Day T. A Biologist’s Guide to Mathematical Modeling in Ecology and Evolution. Princeton: Princeton University Press; 2007.

34. Hurford A, Cownden D, Day T. Next-generation tools for evolutionary invasion analyses. J R Soc Interface. 2010;7: 561–571. doi:10.1098/rsif.2009.0448

35. Ritchie H, Roser M. Age Structure. Our World Data. 2020;

