## Supplemental materials for "Hepatitis B and D: a forecast on actions needed to reduce incidence and achieve elimination"

This appendix provides further details on the hepatitis B and D co-infection model. We outline the differential equations used in model simulations, the methods for estimating the reproductive numbers [16,18] of both hepatitis B and D, and details for the evolutionary invasion analysis [33,34].

### S1 Model equations

**Hepatitis B and D system.** For our mathematical model, we consider a population that divides the population-based on susceptibility to infection, latent infection, chronic infection, and immunity from infection. For hepatitis B, this amounts to susceptible individuals (S), acute hepatitis B infected individuals ($A_B$), chronic hepatitis B infected individuals ($C_B$), the recovered individuals from hepatitis B ($R_B$), and the hepatitis B vaccinated individuals (V). The population infected with hepatitis B is further subdivided based on hepatitis D infection status, namely acute hepatitis B and D infected individuals ($[A_B A_D]$), chronic hepatitis B and acute hepatitis D infected individuals ($[C_B A_D]$), chronic hepatitis B and D infected individuals ($[C_B C_D]$), and chronic hepatitis B infected individuals that have recovered from hepatitis D ($[C_B R_D]$). To describe the transition between compartments for hepatitis B and D, we consider the system of differential equations,

$$\frac{dS}{dt} = b\,N \;\; - b\,\sigma_B \;\; (A_B + C_B) - \lambda_B\,S \; - b\,\sigma_D\,([A_B A_D] + [C_B A_D] + [C_B C_D])$$

$$- (v + \mu)S,$$

$$\frac{dA_B}{dt} = b\,\sigma_B\,(A_B + C_B) + \lambda_B\,S - \gamma_B\,A_B \; - \mu\,A_B - \lambda_D\,A_B,$$

$$\frac{dC_B}{dt} = \rho\,\gamma\,A_B \; - \; (\mu + \mu_B + \delta_B)C_B - \lambda_D\,C_B,$$

$$\frac{dR_B}{dt} = (1-\rho)\gamma_B\,A_B \; + (1-\rho)\gamma_B\,[A_B A_D] + \delta_B\,(C_B + [C_B R_D] + [C_B A_D]) - \mu\,R_B \qquad \text{(S1)}$$

$$+ \delta_D\,([C_B C_D]),$$

$$\frac{d[A_B A_D]}{dt} = b\,\sigma_D\,([A_B A_D] + [C_B A_D] + [C_B C_D]) - \gamma_B\,[A_B A_D] - \mu\,\left[A_{bA_d}\right] + \lambda_D A_B,$$

$$\frac{d[C_B A_D]}{dt} = \rho\,\gamma_B\,[A_B A_D] - \gamma_D\,[C_B A_D] - (\mu + \mu_B + \delta_B)[C_B A_D] + \lambda_D\,C_B,$$

$$\frac{d[C_B C_D]}{dt} = \rho_D\,\gamma_D\,[C_B A_D] - (\mu + \mu_D + \delta_D)[C_B C_D],$$

$$\frac{d[C_B R_D]}{dt} = (1-\rho_D)\gamma_D\,[C_B A_D] \; - (\mu + \mu_B + \delta_B)[C_B R_D],$$

$$\frac{dV}{dt} = v\,S - \mu\,V.$$

where the force of infection for hepatitis B is given by,

$$\lambda_B = \beta_B \frac{A_B + C_B + [A_B A_D] + [C_B A_D] + [C_B C_D] + [C_B R_D]}{N},$$

and the force of infection for hepatitis D is given by,

$$\lambda_D = \beta_D \frac{[A_B A_D] + [C_B A_D] + [C_B C_D]}{N}.$$

**Mutant Hepatitis D system.** If a mutant strain of hepatitis D is permitted, then equations (S1) are augmented to

$$\frac{dS}{dt} = b\,N \; - b\,\sigma_B\;(A_B + C_B) - \lambda_B\,S\; - b\,\sigma_D\,([A_B A_D] + [C_B A_D] + [C_B C_D]) - (v + \mu)S,$$

$$\frac{dA_B}{dt} = b\,\sigma_B\,(A_B + C_B) + \lambda_B\,S - \gamma_B\,A_B\; - \mu\,A_B - (\lambda_D + \lambda_M)\,A_B,$$

$$\frac{dC_B}{dt} = \rho\,\gamma\,A_B\; - \;(\mu + \mu_B + \delta_B)C_B - (\lambda_D + \lambda_M)C_B,$$

$$\frac{dR_B}{dt} = (1-\rho)\gamma_B\,A_B\; + (1-\rho)\gamma_B\,[A_B A_D] + \delta_B\,(C_B + [C_B R_D] + [C_B A_D]) - \mu\,R_B + \delta_D\,([C_B C_D]),$$

$$\frac{d[A_B A_D]}{dt} = b\,\sigma_D\,([A_B A_D] + [C_B A_D] + [C_B C_D]) - \gamma_B\,[A_B A_D] - \mu\,[A_b A_d] + \lambda_D A_B,$$

(S2)

$$\frac{d[C_BA_D]}{dt} = \rho\,\gamma_B\,[A_BA_D] - \gamma_D\,[C_BA_D] - (\mu + \mu_B + \delta_B)[C_BA_D] + \lambda_D\,C_B,$$

$$\frac{d[C_BC_D]}{dt} = \rho_D\,\gamma_D\,[C_BA_D] - (\mu + \mu_D + \delta_D)[C_BC_D],$$

$$\frac{d[C_BR_D]}{dt} = (1 - \rho_D)\gamma_D\,[C_BA_D]\; - (\mu + \mu_B + \delta_B)[C_BR_D],$$

$$\frac{d[A_BA_M]}{dt} = b\,\sigma_D\,([A_BA_M] + [C_BA_M] + [C_BC_M]) - \gamma_B\,[A_BA_M] - \mu\,[A_bA_M] + \lambda_M A_B,$$

$$\frac{d[C_BA_M]}{dt} = \rho\,\gamma_B\,[A_BA_M] - \gamma_M\,[C_BA_M] - (\mu + \mu_B + \delta_B)[C_BA_M] + \lambda_M\,C_B,$$

$$\frac{d[C_BC_M]}{dt} = \rho_M\,\gamma_M\,[C_BA_M] - (\mu + \mu_M + \delta_M)[C_BC_M],$$

$$\frac{d[C_BR_M]}{dt} = (1 - \rho_M)\gamma_M\,[C_BA_M]\; - (\mu + \mu_B + \delta_B)[C_BR_M],$$

$$\frac{dV}{dt} = v\,S - \mu\,V.$$

Here the subscript $M$ denotes infection with the mutant strain of hepatitis D.

Note, for system S2 the force of infection for hepatitis B is given by,

$$\lambda_B = \beta_B\,\frac{A_B + C_B + [A_BA_D] + [C_BA_D] + [C_BC_D] + [C_BR_D] + [A_BA_M] + [C_BA_M] + [C_BC_M] + [C_BR_M]}{N},$$

and the force of infection for the mutant hepatitis D strain is given by,

$$\lambda_M = \beta_M \frac{[A_B A_M] + [C_B A_M] + [C_B C_M] + [C_B R_M]}{N}.$$

**S2 Model equilibria.**

**Hepatitis B and D equilibria.** The model (S1) has 3 valid equilibria: a disease-free equilibrium, a hepatitis B endemic and hepatitis D free equilibrium, and a hepatitis B and D co-endemic equilibrium.

The disease-free equilibrium is

$$S^{DFE} = \frac{b}{\mu + \nu} N,$$

$$V^{DFE} = \frac{\nu}{\mu} \frac{b}{\mu + \nu} N,$$

and

$$A_B^{DFE} = C_B^{DFE} = R_B^{DFE} = [A_B A_D]^{DFE} = [C_B A_D]^{DFE} = [C_B C_D]^{DFE} = [C_B R_D]^{DFE} = 0.$$

When hepatitis B is endemic and hepatitis D free, the equilibrium is

$$\hat{S} = \frac{N}{\beta_B(\mu + \mu_B + \delta_B + \rho_B \sigma_B)} \big(\mu^2 + (\mu_B + \delta_B + \gamma_B - b\sigma_B)\mu + (\mu_B + \delta_B - b\rho_B \sigma_B)\gamma - b\sigma_B(\mu_B + \delta_B)\big),$$

$$\widehat{A_B} = \frac{N}{(\gamma_B + \mu)(\mu + \mu_B + \delta_B + \rho_B\gamma_B)\beta_B}(-\mu^3 - (\mu_B + v + b\sigma_B + \gamma_B + \delta_B)\mu^2$$

$$+ \Big(\big((v + \mu_B + \delta_B + \rho_B\gamma_B)\sigma_B + \beta_B\big)b - (v + \mu_B + \delta_B)\gamma_B - v(\mu_B + \delta_B)\Big)\mu$$

$$+ (\mu_B + \delta_B + \rho_B\gamma_B)(v\sigma_B + \beta_B)b - \gamma v(\mu_B + \delta_B)\big),$$

$$\widehat{C_B} = \frac{\rho_B\gamma_B}{\mu + \mu_B + \delta_B}\widehat{A_B},$$

$$\widehat{R_B} = \frac{(1 - \rho_B)(\mu + \mu_B) + \delta_B}{\rho_B\mu}\,\widehat{C_B},$$

$$\hat{V} = \frac{v}{\mu}\hat{S},$$

and

$$[\widehat{A_B A_D}] = [\widehat{C_B A_D}] = [\widehat{C_B C_D}] = [\widehat{C_B R_D}] = 0.$$

Finally, when both hepatitis B and hepatitis D are endemic, we have the co-endemic equilibrium, which is defined implicitly for the ease of presentation, is

$$\tilde{S} = \frac{bN + b\sigma_B\big(\widetilde{A_B} + \widetilde{C_B}\big) + b\sigma_D\big([\widetilde{A_B A_D}] + [\widetilde{C_B A_D}] + [\widetilde{C_B C_D}]\big)}{\lambda_B + v + \mu},$$

$$\widetilde{A_B} = \lambda_B(\mu + \mu_B + \delta_B + \lambda_D)(\lambda_D^2 + (2\mu + \mu_B + \delta_B + \gamma - b\sigma_B)\lambda_D + \mu^2$$

$$+ (\mu_B + \delta_B + \gamma_B - b\sigma_B)\mu + \gamma_B(\mu_B + \delta_B) - b(\mu_B + \delta_B + \rho_B\gamma_B)\sigma_B)^{-1}\tilde{S}$$

$$\widetilde{C_B} = \frac{\rho_B\gamma_B}{\mu + \mu_B + \delta_B + \lambda_D}\,\widetilde{A_B},$$

$$\widetilde{R_B} = \frac{1}{\mu}\Big((1 - \rho_B)\gamma_B\big(\widetilde{A_B} + [\widetilde{A_B A_D}]\big) + \delta_B\big(\widetilde{C_B} + [\widetilde{C_B A_D}] + [\widetilde{C_B R_D}]\big) + \delta_D[\widetilde{C_B C_D}]\Big),$$

$$[\widetilde{A_B A_D}] = \lambda_D \frac{\psi_1 \mu + \psi_0}{\phi_3 \mu^3 + \phi_2 \mu^2 + \phi_1 \mu + \phi_0},$$

$$[\widetilde{C_B A_D}] = \frac{\rho_B \gamma_B [\widetilde{A_B A_D}] + \lambda_D \widetilde{C_B}}{\mu + \mu_B + \delta_B + \gamma_D},$$

$$[\widetilde{C_B C_D}] = \frac{\rho_D \gamma_D [\widetilde{C_B A_D}]}{\mu + \mu_B + \delta_D},$$

$$[\widetilde{C_B R_D}] = \frac{(1 - \rho_D) \gamma_D [\widetilde{C_B A_D}]}{\mu + \mu_B + \delta_B},$$

$$\tilde{V} = \frac{\nu}{\mu} \tilde{S},$$

where

$\phi_3 = 1,$

$\phi_2 = \mu_D + \delta_B - b\,\sigma_D + \delta_D + \gamma_D + \mu_B + \gamma,$

$$\phi_1 = \big(-(\delta_D + \gamma_D + \rho_B\,\gamma + \mu_D + \mu_B + \delta_B) b\sigma_D + (\mu_B + \gamma + \delta_B + \gamma_D)\mu_D \\ + (\mu_B + \gamma + \delta_B + \gamma_D)\delta_D + \gamma\,(\delta_B + \gamma_D + \mu_B)\big),$$

$$\phi_0 = (-(\delta_B + \gamma_D + p\,\gamma + \mu_B)\mu_D - (\delta_B + \gamma_D + \rho_B\,\gamma + \mu_B)\delta_D - \rho_D\,\gamma_D\,\rho_B\,\gamma) b\,\sigma_D \\ + \gamma\,(\mu_D + \delta_D)\,(\delta_B + \gamma_D + \mu_B),$$

$\psi_1 = b\sigma_D C_B + (\gamma_D + \delta_B + \delta_D + \mu_B + \mu_D) A_B,$

$\psi_0 = (\mu_D + \delta_D)(\delta_B + \gamma_D + \mu_B) A_B + b\,C_B(\rho_D \gamma_D + \delta_D + \mu_D)\sigma_D.$

**S3 The reproductive numbers.**

**The reproductive numbers for Hepatitis B.** To compute the basic reproductive number for hepatitis B we apply the standard next-generation approach. The system (S1) has two stages that contribute to hepatitis B: $A_B$ and $C_B$. By, decoupling the system into new hepatitis B infections and transitions between hepatitis B infection classes, it follows that

$$F_B = \begin{bmatrix} \beta_B \frac{S^{DFE}}{N} + b\sigma_B & \beta_B \frac{S^{DFE}}{N} + b\sigma_B \\ 0 & 0 \end{bmatrix},$$

and

$$V_B = \begin{bmatrix} \gamma_B + \mu & 0 \\ -\rho_B \gamma_B & \delta_B + \mu + \mu_B \end{bmatrix}.$$

Thus, the next-generation matrix is

$$G_B = F_B V_B^{-1} = \begin{bmatrix} N\frac{(\beta_B S^{DFE} + b\sigma_B N)(\mu + \mu_B + \delta_B + \rho_B \gamma_B)}{(\gamma_B + \mu)(\delta_B + \mu + \mu_B)} & \frac{\beta_B S^{DFE} + b\sigma_B N}{N(\delta_B + \mu + \mu_B)} \\ 0 & 0 \end{bmatrix}.$$

From computing the spectral radius of $G_B$, we obtain the basic reproductive number to be

$$\mathcal{R}_0^B = \frac{\beta_B(\mu + \mu_B + \delta_B + \rho_B \gamma_B)}{(\gamma_B + \mu)(\delta_B + \mu + \mu_B)} \frac{S^{DFE}}{N} + \frac{b\sigma_B(\mu + \mu_B + \delta_B + \rho_B \gamma_B)}{(\gamma_B + \mu)(\delta_B + \mu + \mu_B)}, \tag{S3}$$

where $S^{DFE} = \frac{b}{\mu} N$ in the absence of vaccination.

From the calculation of $\mathcal{R}_0^B$, it follows that the effective reproductive number is

$$\mathcal{R}_{eff}^B = \frac{\beta_B(\mu + \mu_B + \delta_B + \rho_B \gamma_B)}{(\gamma_B + \mu)(\delta_B + \mu + \mu_B)} \frac{S}{N} + \frac{b\sigma_B(\mu + \mu_B + \delta_B + \rho_B \gamma_B)}{(\gamma_B + \mu)(\delta_B + \mu + \mu_B)}, \tag{S4}$$

and the control reproductive number is

$$\mathcal{R}_V^B = \frac{\beta_B(\mu + \mu_B + \delta_B + \rho_B\gamma_B)}{(\gamma_B + \mu)(\delta_B + \mu + \mu_B)}\frac{b}{\mu + \nu} + \frac{b\sigma_B(\mu + \mu_B + \delta_B + \rho_B\gamma_B)}{(\gamma_B + \mu)(\delta_B + \mu + \mu_B)}. \tag{S5}$$

**The reproductive numbers for Hepatitis D.** To compute the basic reproductive number for hepatitis D we once again apply the standard next-generation approach. The system (S1) has three stages that contribute to hepatitis D: $[A_B A_D]$, $[C_B A_D]$, and $[C_B C_D]$. By decoupling the system into new hepatitis D infections and transitions between hepatitis D infection classes, it follows that

$$F_D = \begin{bmatrix} \beta_D\frac{\widehat{A_B}}{N} + b\sigma_D & \beta_D\frac{\widehat{A_B}}{N} + b\sigma_D & \beta_D\frac{\widehat{A_B}}{N} + b\sigma_D \\ \beta_D\frac{\widehat{C_B}}{N} & \beta_D\frac{\widehat{C_B}}{N} & \beta_D\frac{\widehat{C_B}}{N} \\ 0 & 0 & 0 \end{bmatrix},$$

and

$$V_D = \begin{bmatrix} \gamma_B + \mu & 0 & 0 \\ -\rho_B\gamma_B & \gamma_D + \delta_B + \mu + \mu_B & 0 \\ 0 & -\rho_D\gamma_D & \delta_D + \mu + \mu_D \end{bmatrix}.$$

Using $F_D$ and $V_D$, we take the spectral radius of $F_D V_D^{-1}$ to estimate the control reproductive of hepatitis D as

$$\mathcal{R}_v^D = \frac{1}{(\gamma_B + \mu)(\gamma_D + \mu + \mu_B + \delta_B)(\mu + \mu_D + \delta_D)}\Big((\mu^2 + (\mu_B + \mu_D + \rho\gamma_B + \gamma_D + \delta_B + \delta_D)\mu + (\mu_B + \gamma_D + \delta_B + \rho\gamma_B)(\mu_D + \delta_D) + \rho\gamma\rho_D\gamma_D)\left(\beta_D\frac{\widehat{A_B}}{N} + b\sigma_D\right) + \beta_D(\gamma_B + \mu)(\rho_D\gamma_D + \mu + \mu_D + \delta_D)\frac{\widehat{C_B}}{N}\Big). \tag{S6}$$

$$\mathcal{R}_v^D = \frac{1}{(\gamma_B+\mu)(\gamma_D+\mu+\mu_B+\delta_B)(\mu+\mu_D+\delta_D)}\Bigg(\big(\mu^2+(\mu_B+\mu_D+\rho\gamma_B+\gamma_D+\delta_B+\delta_D)\mu + (\mu_B+\gamma_D+\delta_B+\rho\gamma_B)(\mu_D+\delta_D)+\rho\gamma\rho_D\gamma_D\big)\left(\beta_D\frac{\widehat{A_B}}{N} + b\sigma_D\right) + \beta_D(\gamma_B+\mu)(\rho_D\gamma_D+\mu+\mu_D+\delta_D)\frac{\rho_B\gamma_B}{\mu+\mu_B+\delta_B}\frac{\widehat{A_B}}{N}\Bigg)$$

Finally, the basic reproductive number is obtained by

$$\mathcal{R}_0^D = \mathcal{R}_v^D|_{v=0}. \tag{S7}$$

**The mutant reproductive number for Hepatitis D.** To determine if a more virulent strain of hepatitis D can invade the population we follow the approach from the next-generation method [34]. Following this approach, we estimate the reproductive number evaluated at hepatitis B and D co-endemic equilibrium, where $[A_B A_M] = [C_B A_M] = [C_B C_M] = 0$. To do this, we decompose system (S2) into matrices $F_{ext}$ and $V_{ext}$:

$$F_{ext} = \begin{bmatrix} \frac{\beta_D\,\widetilde{A_B}}{N}+b\sigma_D & \frac{\beta_D\,\widetilde{A_B}}{N}+b\sigma_D & \frac{\beta_D\,\widetilde{A_B}}{N}+b\sigma_D & 0 & 0 & 0 \\ \beta_D\frac{\widehat{C_B}}{N} & \beta_D\frac{\widehat{C_B}}{N} & \beta_D\frac{\widehat{C_B}}{N} & 0 & 0 & 0 \\ 0 & 0 & 0 & 0 & 0 & 0 \\ 0 & 0 & 0 & \frac{\beta_M\,\widetilde{A_B}}{N} & \frac{\beta_M\,\widetilde{A_B}}{N} & \frac{\beta_M\,\widetilde{A_B}}{N} \\ 0 & 0 & 0 & \frac{\beta_M\,\widetilde{C_B}}{N} & \frac{\beta_M\,\widetilde{C_B}}{N} & \frac{\beta_M\,\widetilde{C_B}}{N} \\ 0 & 0 & 0 & 0 & 0 & 0 \end{bmatrix}$$

and

$$V_{ext} = \begin{bmatrix} \gamma_B + \mu & 0 & 0 & 0 & 0 & 0 \\ -\rho_B\gamma_B & \gamma_D + \delta_B + \mu + \mu_B & 0 & 0 & 0 & 0 \\ 0 & -\rho_D\gamma_D & \delta_D + \mu + \mu_D & 0 & 0 & 0 \\ 0 & 0 & 0 & \gamma_B + \mu & 0 & 0 \\ 0 & 0 & 0 & -\rho_B\gamma_B & \gamma_M + \delta_B + \mu + \mu_B & 0 \\ 0 & 0 & 0 & 0 & -\rho_M\gamma_M & \mu + \mu_B + \mu_D \end{bmatrix}.$$

Next, we determine the next-generation matrix

$$G_{ext} = F_{ext}V_{ext}^{-1} = \begin{bmatrix} F_D V_D^{-1} & \mathbf{0}_{3\times 3} \\ \mathbf{0}_{3\times 3} & F_M V_M^{-1} \end{bmatrix}.$$

The mutant reproductive number corresponds to $\rho(F_M V_M^{-1})$, and is given by

$$\mathcal{R}_M^D = \frac{\beta_M(\mu^2 + (\mu_B + \mu_M + \rho\gamma_B + \gamma_M + \delta_B + \delta_M)\mu + (\mu_B + \gamma_M + \delta_B + \rho\gamma_B)(\mu_M + \delta_M) + \rho_B\gamma_B\rho_M\gamma_M)}{(\gamma_B + \mu)(\gamma_M + \mu + \mu_B + \delta_B)(\mu + \mu_M + \delta_M)}\frac{\widetilde{A_B}}{N} + \frac{\beta_M(\mu + \mu_M + \delta_M + \rho_M\gamma_M\ )}{(\gamma_M + \mu + \mu_B + \delta_B)(\mu + \mu_M + \delta_M)}\frac{\widetilde{C_B}}{N}, \quad \text{(S8)}$$

and by the competitive exclusion principle, the invasion of the mutant strain can occur provided

$$\mathcal{R}_M^D > \mathcal{R}_0^D. \quad \text{(S9)}$$

**S4 The transmission and vaccination rates.** To estimate the transmission rate of hepatitis b we use estimates on the prevalence of hepatitis B, the proportion of chronic hepatitis B, the basic reproductive number of hepatitis B [11] along with the endemic equilibrium for hepatitis B (S3). It follows for $\mathcal{R}_0^B = 2.406$ [11] that

$$\beta_B \approx 0.391 \text{ year}^{-1}.$$

Chronic infection with hepatitis B reduces the average life span of 78.8 years by approximately 4.8 years [28]. Thus, on average, we have that

$$\frac{1}{\mu + \mu_B} \approx 74 \text{ years}.$$

It follows that $\mu_B \approx 0.00086 \text{ year}^{-1}$. Similarly, chronic hepatitis D co-infection typically leads to death 10 years sooner than mono-infection with hepatitis [29]. Thus, we have that

$$\frac{1}{\mu + \mu_D} \approx 60.8 \text{ years},$$

and so

$$\mu_D \approx 0.003 \text{ year}^{-1}$$

To estimate the transmission rate of hepatitis D, estimates place $\mathcal{R}_v^D \approx 1.01$ with an $\mathcal{R}_0^D \approx 1.41$ under an average lifespan of 55.6 years [20]. When considering a lifespan of $\frac{1}{\mu}$ =78.8 years, we have that $\mathcal{R}_v^D \approx 2.87$ when $v = 0.00025 \text{ year}^{-1}$ . Given this estimate, and the parameter values from Table 1, it follows from (S6) that

$$\beta_B \approx 25.76 \text{ year}^{-1},$$

and in the absence of vaccination that

$$\mathcal{R}_0^D \approx 3.33.$$

**The proportion that develops active disease.** To estimate the proportion of individuals that develop active disease we use current estimates that 90% of perinatal infections are active,

30% of childhood infections (<6 years) are active, and 10% of other infections are active [28]. Thus, given that approximately 0.142% of individuals are classified as perinatal, 2.9% are classified as children under 6 years [35], it follows that

$$\rho = 0.9(0.00142) + 0.3(0.028) + 0.1(0.96.9) \approx 0.101.$$

### References


1. Jefferies M, Rauff B, Rashid H, Lam T, Rafiq S. Update on global epidemiology of viral hepatitis and preventive strategies. World J Clin cases. 2018;6: 589–599. doi:10.12998/wjcc.v6.i13.589
